# Severity of SARS-CoV-2 reinfections in second wave determines likelihood of mild endemicity

**DOI:** 10.1101/2021.07.21.21260944

**Authors:** Jennie S Lavine, Ottar N Bjornstad, Daniel Coombs, Rustom Antia

## Abstract

Immunity to SARS-CoV-2 is building up globally, but will this be sufficient to prevent future COVID-19 epidemics in the face of variants and waning immunity? Manaus, Brazil offers a concerning glimpse of what may come: six months after the majority of the city’s population experienced primary infection, a second wave with a new strain resulted in more deaths than the first wave. Current hypotheses for this surge rely on prior immunity waning due to time and antigenic distance. Here we show this hypothesis predicts a severe endemic state. We propose an alternative hypothesis in which individuals infected in the first wave lose protection against transmission but retain immunity against severe disease and show this hypothesis is equally compatible with existing data. In this scenario, the increased number of deaths is due to an increased infection fatality ratio (IFR) for primary infections with the new variant. This alternative predicts a mild endemic state will be reached within decades. Collecting data on the severity of reinfections and infections post-vaccination as a function of time and antigenic distance from the original exposure is crucial for optimizing control strategies.

---

The severe second wave of COVID-related deaths in Manaus, Brazil following an already very large first wave (Fig 1 gray dots) raises concerns of deadly reinfections with SARS-CoV-2. The first wave is thought to have receded at least in part due to widespread immunity [1]. Current hypotheses for this large and unexpected outbreak that appears to have started in early December, 2020 suggest a critical role for higher transmissibility and virulence of the P.1 variant and loss of immunity of previously infected hosts due to waning and antigenic changes giving rise to deadly second infections [2, 3, 4]. This scenario suggests immunity generated by early SARS-CoV-2 strains (and likely vaccines) provide little protection against reinfection with new more dominant variants and will require reformulation of current policies regarding vaccination, relaxing NPIs, and other public health measures. Here we suggest a second scenario with different long-term consequences which we show is also fully consistent current data from Manaus.

**Figure 1:**
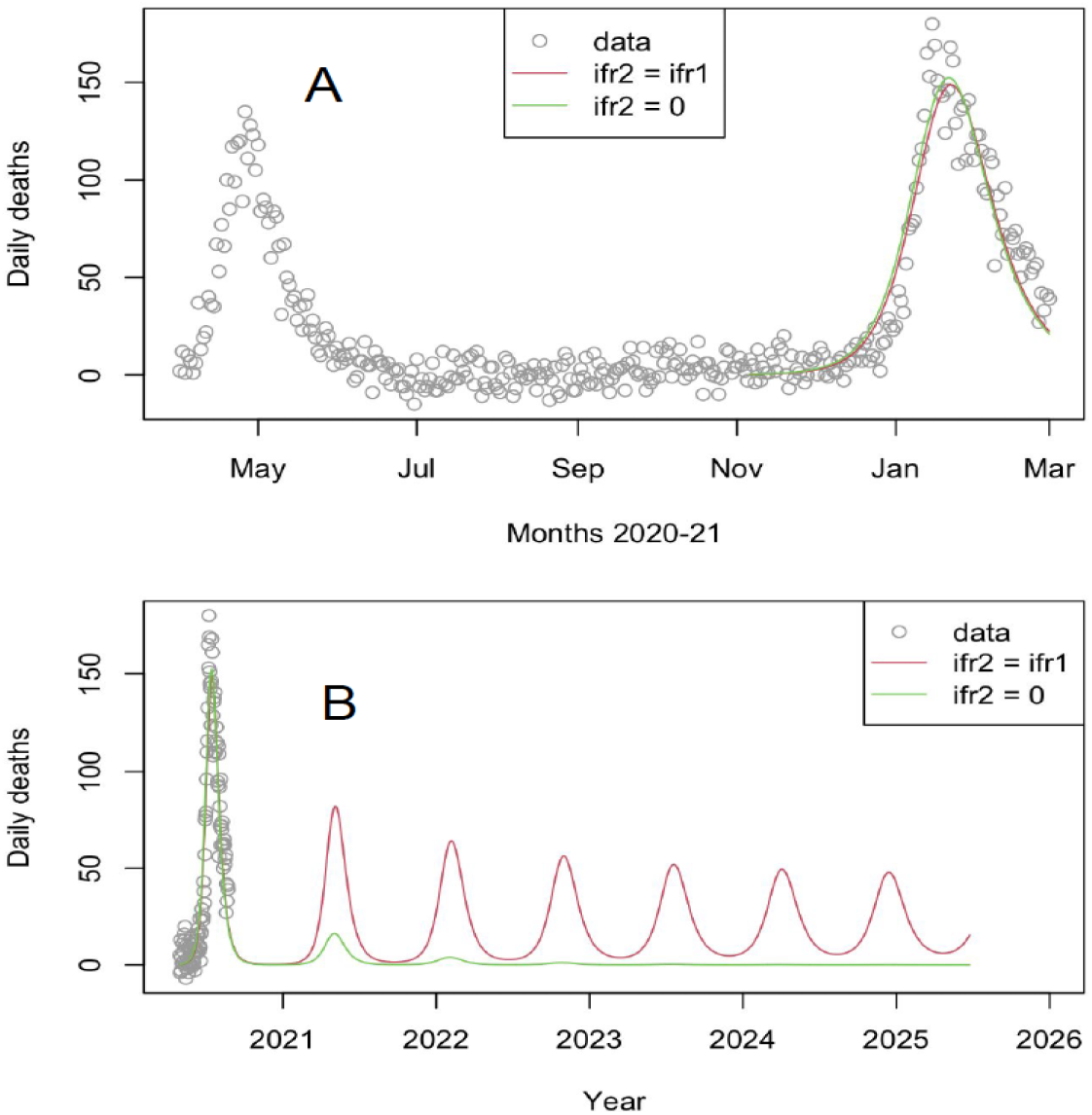
Mortality during the second wave is equally compatible with models in which deaths stem only from primary infections or a mix of primary and secondary infections (neither model’s AIC weight > 0.95) but the long-term consequences differ. Daily excess deaths in Manaus are plotted from the start of the first wave to March 2021 (gray dots). Best fit model simulations (lines) are shown following the introduction of the P.1 strain. In both scenarios, the best fit R0 = 3 and mean duration of transmission-blocking immunity = 0.4 yr. In scenario 1 (red curve), prior infection does not protect against pathology in the second wave and infections in the second wave (mostly with the P.1 strain) are fit to be 1.7 times as fatal as those in the first wave and 70% of deaths occur in secondary cases. In scenario 2 (green curve), immunity generated by infection during the first wave protects against death following infection during the second wave (secondary IFR=0), so all deaths occur in primary infections and second wave infections are fit at 5.7 times as fatal as first wave. The long-term consequences of the two scenarios differ widely. In scenario 1 (red line), transmission-blocking and disease-blocking immunity are lost concurrently and the model predicts large severe outbreaks many years into the future. In contrast, in scenario 2 (green line) a prior infection protects against death though not transmission, and a mild endemic state is reached within a year or two.

Recent studies demonstrate we need a nuanced understanding to describe adaptive immunity to SARS-CoV-2 [5]. Current data indicate that T cell immunity is protective across a wider range of variants [6] and, while neutralizing antibodies have reduced efficacy against the new P.1 variant (the dominant strain in Manaus in the second wave) [7], there is no significant difference in T-cell-based immunity in its reactivity to this strain [8]. There is a paucity of data on the effect of primary exposure and partial immunity that may reduce the severity of reinfection (in SARS-CoV-2 or any other pathogen for that matter), but evidence from experimental infection studies with the endemic coronaviruses and SARS-CoV-2 vaccine trials suggests that lack of transmission-blocking immunity leaves people susceptible to reinfection but with less severe pathology [9, 10].

We therefore postulate a second scenario in which mildly symptomatic transmissible reinfections are adding fuel to the second wave in Manaus but deaths arise predominantly from primary infections. Here, we use a model of SARS-CoV-2 transmission and disease that incorporates recent immunological observations regarding the ‘dimmer switch’ nature of coronavirus immunity ([11], equations in SI) to model the two scenarios described above and find they explain the data equally well (Fig 1 red for the first scenario and green for the second) (diff in log like). While the simulations of the two scenarios are indistinguishable for the second wave, their projections have very different predictions for disease burden in the coming years (Fig 1b). In the first scenario, loss of immunity due to a combination of waning and virus evolution predicts a severe endemic state. In contrast, in the second scenario (a single infection protects against severe disease upon reinfection with new strains), the prediction is of a mild endemic state. These predictions are qualitatively robust to the precise level of population immunity following the first wave (Fig 2).

**Figure 2:**
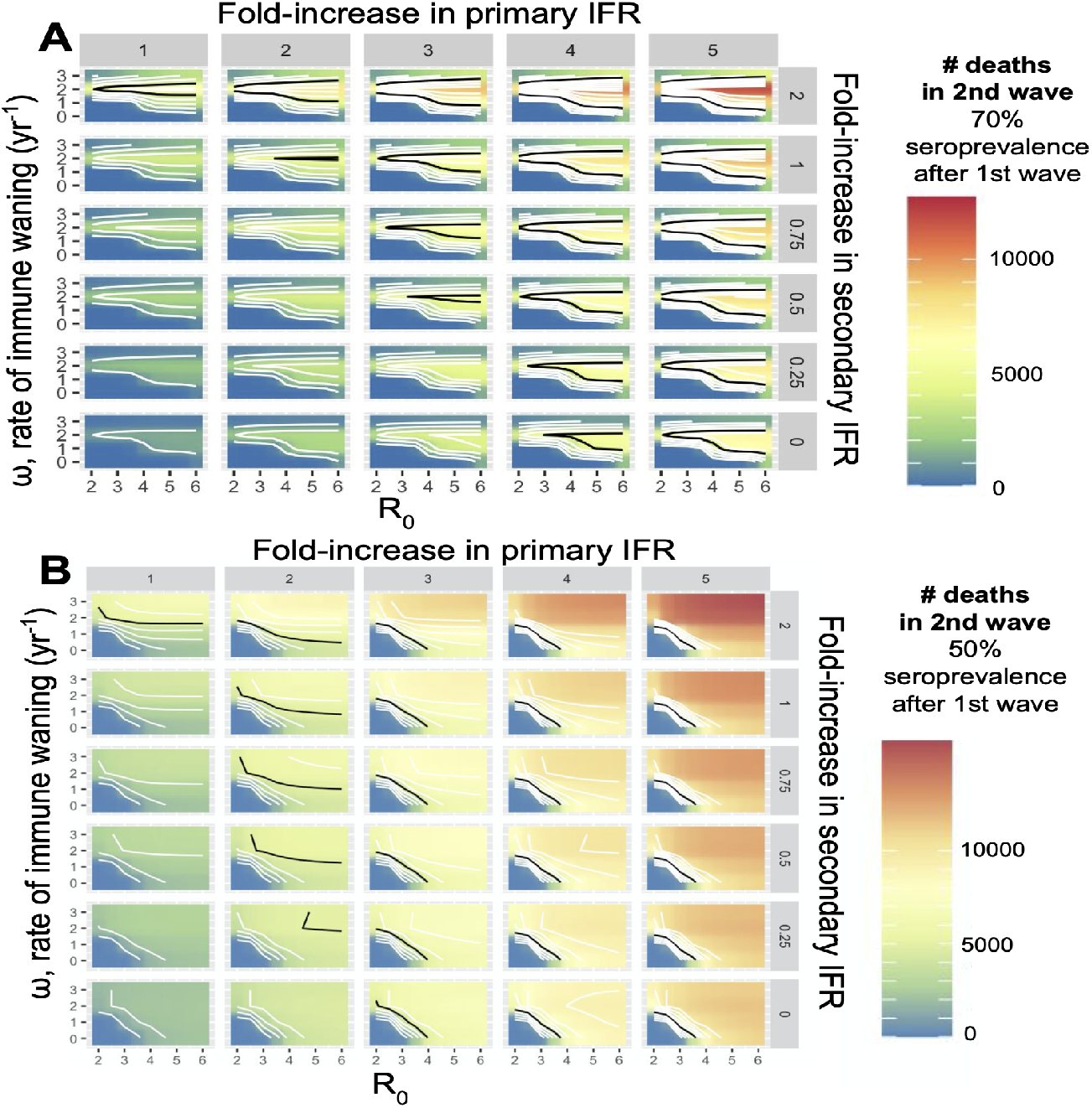
There is a trade-off between the IFR of primary and secondary infections to explain the total deaths in the second wave (black lines). The predicted number of deaths during the second wave is impacted by the duration of immunity (*ω*, small y-axes), *R*_0_ (small x-axes), and the fold change in the primary and secondary IFRs (meta x- and y-axes, respectively). The heat maps show the predicted number of deaths during the second wave for combinations of these parameters. Panel **A** shows results assuming 70% of the population immune at the end of the first wave. The upper black lines are consistent with rapidly waning immunity, the lower black lines with slower waning. Panel **B** assumes only 50% of the population was immune at the end of the first wave. The *R*_0_ for the pre-P.1 infections is set to two for consistency with the observed lack of infections or excess deaths in June-Oct. The number of deaths in the second wave is less consistent with very rapid waning assuming this higher level of susceptibility following the first wave. It would generate more deaths than observed (bright red in upper right corners, no upper black lines). Nonetheless, a trade-off is observed between the IFR of primary and secondary infections for explaining the excess death data with longer lasting immunity, highlighting the same need for collecting data on the severity of reinfections

Consideration of these alternative scenarios is likely to be relevant for policy making following vaccination as well as infection. Understanding how vaccine-induced immunity differs from infection-induced immunity will be crucial. Current evidence suggests virus- and vaccine-elicited immunity may differ in magnitude (higher antibody titers to vaccine), breadth (vaccine typically focused on spike protein) and localization (systemic vs respiratory tract) [12].

Our mathematical model of the two scenarios suggests that aggregated data on infections and disease outcomes alone do not contain sufficient information to determine the contribution of primary and secondary infections to disease burden. It is therefore crucial to measure how disease severity (IFRs) and virus transmission change as a function of immune status and antigenic evolution.

Retrospective studies to connect individual health records of cases and deaths with prior serostatus or infection status [13] are underway, and similar protocols may be useful in other regions, including those with high vaccine coverage (e.g., Israel [14]) and variants of concern [15]. Ideally, this would be done in conjunction with prospective longitudinal studies that integrate detailed measurements of IgG and IgA titers and T-cell responses [16, 17] while monitoring for infection and disease severity in naive, vaccinated and naturally infected cohorts. Such studies would allow us to distinguish whether the resurgence of cases is due to rapid waning of disease-reducing immunity or severe disease in immunologically naive individuals. This distinction is crucial in determining whether the combination of vaccination and widespread circulation of the virus will result in a mild endemic state, or whether frequent re-formulation of the vaccine, potentially in combination with the development of effective antiviral treatment strategies and further NPIs, will be needed in the months or years ahead.

## Materials and Methods

### Data

Death data from Manaus during the pandemic are based on cemetery and crematorium records in the city of Manaus.

Baseline deaths are calculated as a 15-day running average of adjusted deaths from 2019 https://www.google.com/url?q=https://github.com/capyvara&sa=D&source=editors&ust=1616610609031000&usg=AFQjCNHxIwl_gKvzB1sTJ-WO6Lk-5Z0BFw.

Excess deaths are calculated by subtracting the running average in 2019 from the same date during the pandemic https://docs.google.com/spreadsheets/d/1UMhKX4mBSBM8YCZL338vB0UY8TYsgGTb/edit#gid=1271932269.

The birth rate is taken from https://g1.globo.com/am/amazonas/noticia/2020/12/11/amazonas-lidera-ranking-de-registros-de-nascimento-tardios-no-pais-aponta-ibge.ghtml.

The age distribution of the population in Manaus is taken from https://www.citypopulation.de/en/brazil/amazonas/manaus/130260305__manaus/.

The background age-specific death rate is assumed to be the same for Manaus as for the state of Amazonas as a whole in 2019 and is taken from https://transparencia.registrocivil.org.br/dados-covid-download.

The age-specific infection fatality ratio is taken from the supplement of Buss et al.

### Model

The model is an extension of Lavine et al [11] (see SI for equations). The model assumes that after one infection, there is a refractory period representing sterilizing immunity. The duration of the refractory period is Gamma-distributed. Following that, individuals are susceptible to reinfection. For the purpose of scenario analysis, the basic model assumes that reinfections are half as transmissible as primary infections but reinfecteds may be protected against disease. In the SI, we consider partial protection against disease (*IFR*1 > *IFR*2 > 0) and an accumulation of protection over the course of two infections (*IFR*1 > *IFR*2 > *IFR*3). We additionally show that the results are not sensitive to higher transmissibility of reinfections.

For Manaus, it is plausible that 70% of the population was was infected in the intense first wave, during April and May, 2020. We therefore assume 70% of the population acquired immunity in May 2020 (approximately half way through the first wave). We simulate from these equations with a basic reproduction number, *R*_0_ = 3 (the high end of the R0 estimated during wave 1 in Manaus) [1] from May 1 - Nov 6, the date on which the P.1 strain was estimated to have emerged [3]. Our model recapitulates the low number of cases throughout the summer and early fall and tracks the decay of immunity during this time. On Nov 6, the simulation for the fit is started as per the previous trajectory to model the second wave. Daily deaths are calculated from the simulated dynamics and compared with the data.

### Statistics

The simulations (Nov 6, 2020 - Mar 1, 2021) are fit to the death data from the second wave using maximum likelihood estimation ([18, 19]). The error around the data is assumed to be normally distributed and we estimate the IFR (a multiplier of the IFR from the first wave), the mean duration of the refractory period, and R0. The other parameters are fixed as follows: Gamma shape parameter = 9, infectious period = 9 days, and secondary infections are half as transmissible as primary infections (rho=0.5).

All code is available on GitHub (https://doi.org/10.5281/zenodo.4666527).

## Data Availability

All code and data is linked to on GitHub and archived on Zenodo.

https://doi.org/10.5281/zenodo.4666527

## Funding

This work was funded by NIH grants U01 AI150747, U01 HL139483 and U01 AI144616

Data and materials availability: The scripts and data used to perform the analysis and generate the figures in this paper are available on GitHub (http://github.com/JennieLavine/secondary_ifr) and archived in Zenodo (https://doi.org/10.5281/zenodo.4666527).

## Supplementary Information

This model is simulated from the following equations, where *j* represents the age class and *k* the particular compartment of a Gamma chain:

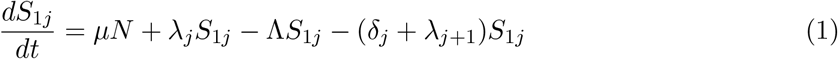

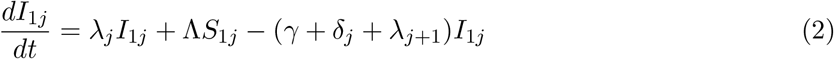

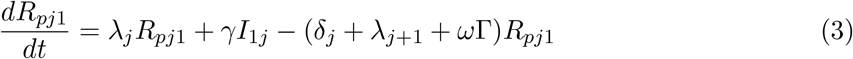

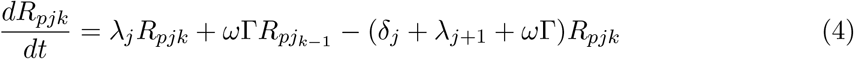

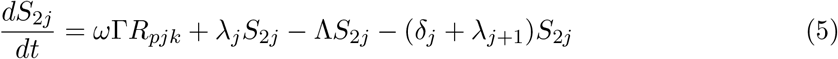

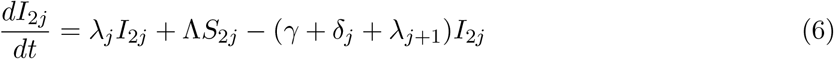

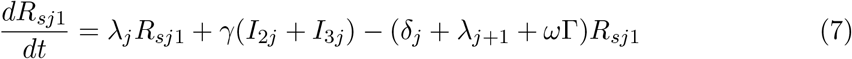

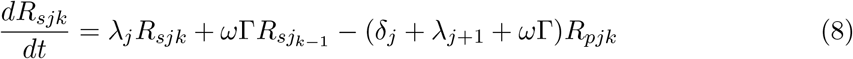

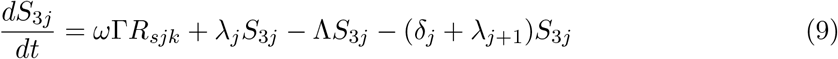

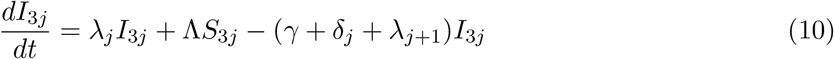

The force of infection is therefore calculated according to

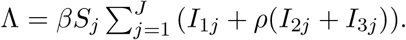

The Gamma-distributed duration of immunity (*R*) is modeled by a chain of Γ *R*-compartments for each *R* class.

If one infection provides partial protection against subsequent disease, it may be that a second infection will lead to increased protection (similar to a second dose of vaccine). If that is the case, even if the secondary IFR is relatively high (but lower than the primary IFR), the long-term outcome may still be rather optimistic (Fig S1).

**Figure 3: Supp Fig 1:**
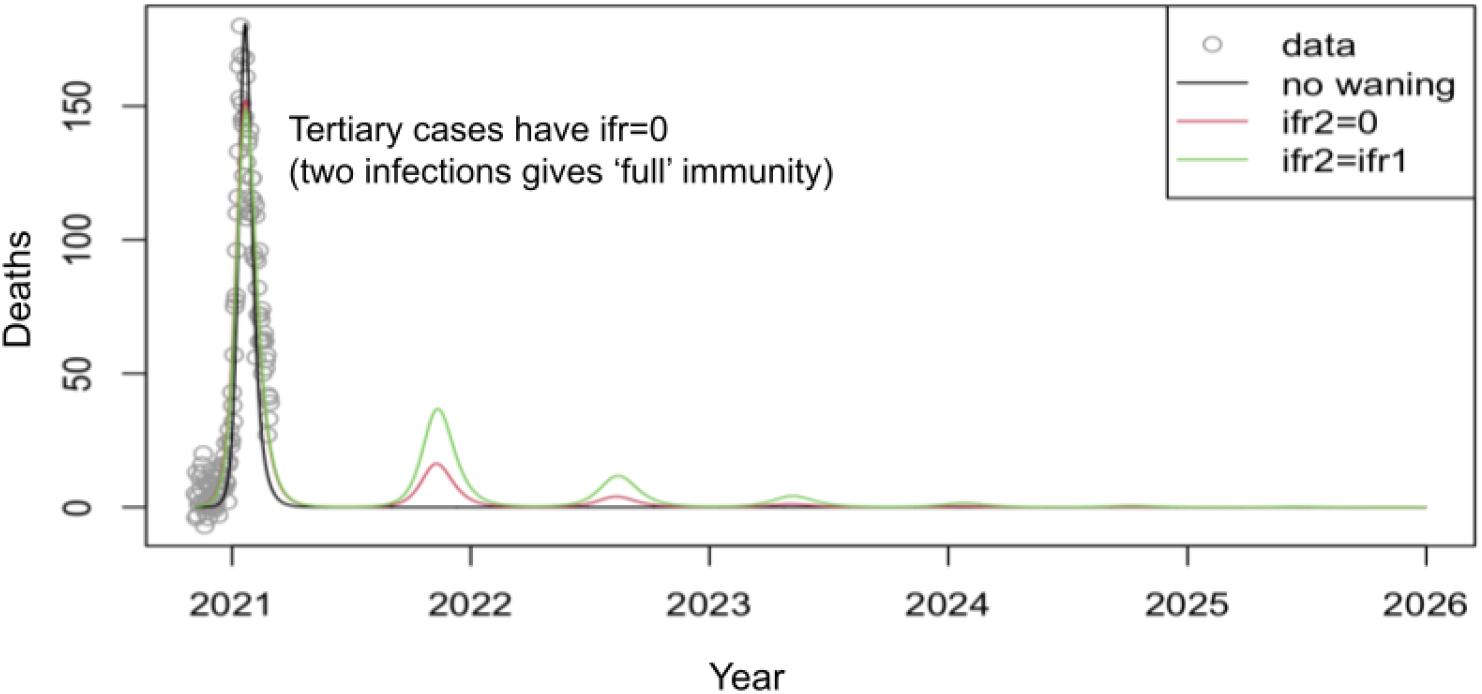
A long-term mild endemic state is predicted when protection against severe disease is acquired across multiple infections. Here, the severity of secondary infections are as shown in the fits in the main text, but people getting tertiary and subsequent infections are assumed to be protected.

